# Characteristics of patients with COVID-19 during epidemic ongoing outbreak in Wuhan, China

**DOI:** 10.1101/2020.03.19.20033175

**Authors:** Xiaomin Luo, Hongxia Xia, Weize Yang, Benchao Wang, Tangxi Guo, Jun Xiong, Zongping Jiang, Yu Liu, Xiaojie Yan, W Zhou, Lu Ye, Bicheng Zhang

**Affiliations:** Department of Emergency, Eastern Campus, Renmin hospital of Wuhan University, Wuhan, China; Cancer center, Renmin hospital of Wuhan University, Wuhan, China

**Keywords:** COVID-2019, SARS-CoV-2, homeostasis disturbance, inflammatory response, early intervention

## Abstract

**Background:** Since Dec 2019, SARS-CoV-2 has caused about fifty thousand patients and over two thousand deaths in Wuhan, China. We reported characteristics of patients with COVID-19 during epidemic ongoing outbreak in Wuhan.

**Methods:** Data of COVID-19 patients with clinical outcome in a designated hospital in Wuhan, were retrospectively collected from electronic medical records. Characteristics were compared between patients who died or recovered, and between patients with different disease severity.

**Results:** By Feb 25, 2020, 403 patients were enrolled, 100 died and 303 recovered. Most of non-survivors tended to be males, old aged, or with chronic diseases. Duration from illness onset to admission was 9 (7-12) days. Patients with severe or critical illness had more days from onset to admission compared to those with ordinary illness. Lymphopenia, anemia, hypoproteinemia, and abnormal serum sodium were presented in 52.6%, 54.6%, 69.8%, and 21.8% cases, respectively. Patients who died or with severe/critical illness showed increased white blood cell and neutrophil count, serum total bilirubin, creatinine, hypersensitive troponin I, D-dimer, procalcitonin, and C-reactive protein, and decreased red blood cell, lymphocyte, platelet count, and serum albumin on admission compared to those who recovered or with ordinary illness. Complications of acute organ injury and secondary infection were common in patients with COVID-19, especially in non-survivors.

**Conclusions:** Multiple homeostasis disturbances were common in patients with severe or critical illness at admission. Early support should be provided, especially for old men with chronic disease, which is vital to control disease progression and reduce mortality of COVID-19 during epidemic ongoing outbreak.

## Introduction

In late December 2019, cluster cases of pneumonia of unknown etiology were reported by local health facilities in Wuhan, China.^1^ A novel coronavirus was identified as the pathogen of this acute respiratory illness, which was named SARS-Cov-2 by World Health Organization (WHO) for its similarity in gene sequence to severe acute respiratory syndrome coronavirus (SARS-CoV).^2,3^ The disease caused by SARS-Cov-2 was latterly designated COVID-19.^4^ The SARS-CoV-2 epidemic has rapidly spread from Wuhan to other areas. On 30 Jan 2020, WHO declared SARS-CoV-2 epidemic a public health emergency of international concern.^3^ As a newly discovered virus, SARS-CoV-2 has brought about much more serious global health threats than the two former major pandemics by other members of coronavirus family, SARS-CoV and Middle East respiratory syndrome coronavirus (MERS-CoV).^5^ As of Mar 6, 2020, a total of 98192 cases of COVID-19 have been confirmed globally, including 80711cases in China and 17481 cases in other 88 countries/territories/areas. Of them, 3380 patients died, 3045 in China and 335 in abroad.^6^

Data from the early epidemic revealed that common symptoms of SARS-CoV-2 infection included fever, fatigue, dry cough, and shortness of breath, and older males with comorbidities are more susceptible to the SARS-CoV-2 infection and progressive to severe and even fatal respiratory diseases.^7,8^ Human-to-human transmission has been confirmed mainly through respiratory droplets and close contact.^8,9^ The management of patients with SARS-CoV-2 infection is mainly symptomatic treatment and life support for severely and critically ill patients. Anti-viral treatment has been listed in protocols issued by the Chinese National Health Commission (NHC), however, these antiviral agents have yet to be confirmed effective for SARS-CoV-2.^9^ Currently, SARS-CoV-2 has infected most patients and caused most deaths in Wuhan, China. However, little information focusing on final outcome and disease severity on admission can be obtained associated with clinical characteristics during the ongoing outbreak of SARS-CoV-2 pneumonia. In this retrospective study, we aimed to describe and compare the clinical features of patients with COVID-19 related to final outcome and disease severity on admission during its ongoing outbreak in Wuhan, China.

## Materials and methods

### Study design and participants

For this retrospective, single-center study, patients who admitted to Eastern Campus of Renmin Hospital, Wuhan University, were recruited from Jan 30 to Feb 25, 2020. Renmin Hospital of Wuhan University is a teaching hospital affiliated to Wuhan University in Wuhan, China. With epidemic ongoing outbreak, Eastern Campus of our hospital was requisitioned as one of designated hospitals for SARS-CoV-2 infection on Jan 25, 2020. After being transformed into an infectious disease hospital, Eastern Campus began to admit patients who were diagnosed as COVID-19 according to protocol^9^ on Jan 30, 2020. Clinical outcome of all patients was reviewed till Feb 25, 2020. Patients who were still hospitalized by that time were excluded in this study. This study was approved by the Research Ethics Committee of Renmin Hospital of Wuhan University (approval number: WDRM 2020-K038). Written consent was not required for patients since it was a retrospective, observational study.

### Data collection

Information of each patient was obtained mainly through screening Electronic Health Records and Laboratory Information Management System supplied by DHC Software Co., Ltd (Beijing, China). Nursing records were also reviewed if necessary. Patients’ demographic information, history of smoking or drinking, chronic diseases, and onset symptoms, were collected, as well as duration from illness onset to hospital admission and disease severity on admission. Indexes of laboratory test on admission were evaluated, including complete blood count, Clreactive protein (CRP), blood biochemistry, myocardial enzymes, and coagulating function. Findings of chest computed tomographic scan (CT) or x-ray film on admission were reviewed and listed. Disease severity (ordinary, severe, or critical) was determined according to the guidance issued by NHC^9^. Treatment measures evaluated in this research contained high-flow nasal cannula oxygen therapy (HFNC), non-invasive mechanical ventilation (NIMV) or invasive mechanical ventilation (IMV), blood purification treatment, application of antiviral agents, antibacterial agents, anti-fungal agents, corticosteroids, and immunoglobulin. The primary endpoint of this study was patients’ clinical outcome (died or recovered), and secondary endpoint was disease severity on admission. Criteria for recovery were as follows: temperature returned to normal for more than 3 days, respiratory symptoms improved significantly, and negative results of two consecutive SARS-CoV-2 RNA detection in respiratory samples (sampling interval at least 1 day).^9^

### Statistical analysis

Categorical variables were presented as number (%), and continuous variables were described as mean (standard deviation, SD) for normally distributed data or median (interquartile range, IQR) for abnormally distributed data. Independent t test was used to compare continuous variables with normal distribution; otherwise, the Mann-Whitney test was used. Chi-squared test was conducted to compare categorical variables. All statistical analyses were performed using IBM SPSS Statistics 19.0 (SPSS Inc.). A two-sided test of α < 0.05 was considered statistical significance.

## Results

By Feb 25, 2020, 985 patients with confirmed COVID-19 were admitted to Eastern Campus of Renmin Hospital of Wuhan University. Five hundred and seventy four patients were still in hospitalization at that time. Four hundred and eleven patients had clinical outcome, among them 8 died on admission day. Consequently, 403 cases were finally enrolled in this study, 100 died and 303 recovered (figure 1). The median age was 56 (39–68) years, 193 (47.9%) were males. Non-survivors had a significantly older age than survivors (median 71 (65–80) years vs. 49 (37–62) years, p<0.001). The mortality was obviously higher in males than in females. Twenty nine (7.2%) patients had history of smoking, and 41 (10.2%) had history of drinking. History of smoking or drinking was not associated with clinical outcome. One hundred and seventy five (43.4%) patients had chronic diseases. Non-survivors had a higher incidence of chronic diseases compared to survivors (77.0% vs. 32.3%, p<0.001). One hundred and thirteen (28%) patients had hypertension. Thirty nine (9.7%) patients had cerebrovascular diseases. Fifty seven (14.1%) patients had diabetes. Besides, coronary heart disease, chronic pulmonary disease, cirrhosis, anemia, and chronic kidney disease appeared in 36 (8.9%), 28 (6.9%), 25 (6.2%), 15 (3.7%), and 7 (1.7%) patients, respectively. Fever accounted for the most main symptom of onset (79.2%).

**Figure 1.**
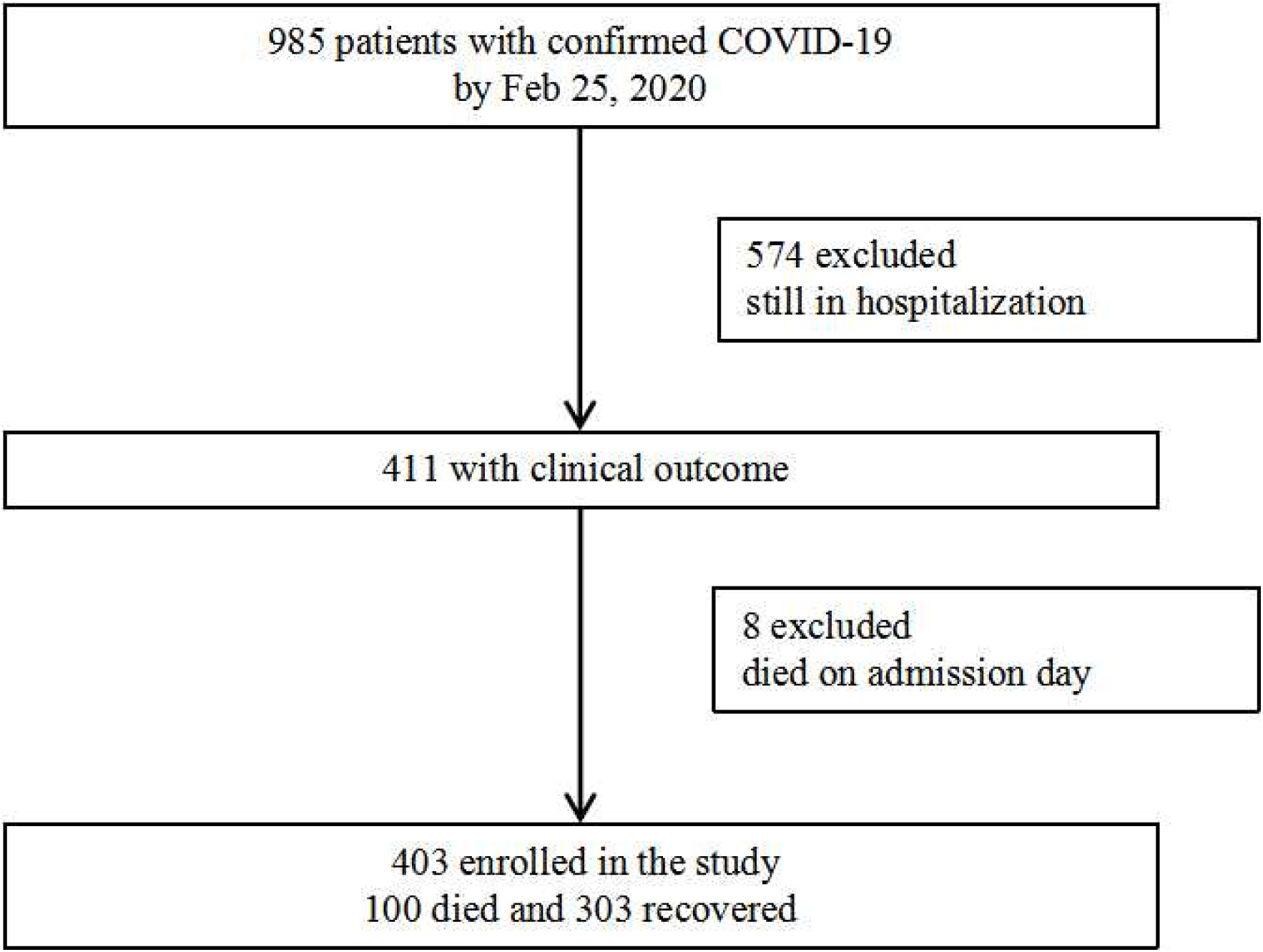
study flow char.

Other main onset symptoms were cough (10.2%) and fatigue (3.5%). The median duration from onset of symptoms to hospital admission was 9 (7–12) days. On admission, all patients had abnormal lung image manifested by chest computed tomography (CT) scan or x-ray film. The typical abnormality in chest CT image was bilateral ground glass opacity and subsegmental consolidation. Bilateral involvement appeared in 376 (93.3%) patients (table 1).

**Table 1.** Demographic and epidemicological characteristics of COVID-19 patients.

|  | All patients (n=403) | Died (n=100) | Recovered (n=303) | P |
| --- | --- | --- | --- | --- |
| Age, years | 56 (39-68) | 71 (65-80) | 49 (37-62) | 0.000 |
| < 60 | 232 (57.6%) | 20 (20%) | 212 (70.0%) | 0.000 |
| ≥ 60 | 171 (42.4%) | 80 (80%) | 91 (30.0%) |  |
| Gender |  |  |  | 0.035 |
| Male | 193 (47.9%) | 57 (57%) | 136 (44.9%) |  |
| Female | 210 (52.1%) | 43 (43%) | 167 (55.1%) |  |
| Smoking | 29 (7.2%) | 9 (9%) | 20 (6.6%) | 0.421 |
| Drinking | 41 (10.2%) | 10 (10%) | 31 (10.2%) | 0.947 |
| With chronic diseases | 175 (43.4%) | 77 (77%) | 98 (32.3%) | 0.000 |
| Hypertension | 113 (28.0%) | 60 (60%) | 53 (17.5%) | 0.000 |
| Cerebrovascular diseases | 39 (9.7%) | 22 (22%) | 17 (5.6%) | 0.000 |
| Coronary heart disease | 36 (8.9%) | 16 (16%) | 20 (6.6%) | 0.004 |
| Diabetes | 57 (14.1%) | 25 (25%) | 32 (10.6%) | 0.000 |
| Chronic pulmonary disease | 28 (6.9%) | 17 (17%) | 11 (3.6%) | 0.000 |
| Cirrhosis | 25 (6.2%) | 9 (9%) | 16 (5.3%) | 0.181 |
| Chronic kidney disease | 7 (1.7%) | 3 (3%) | 4 (1.3%) | 0.501 |
| Anemia | 15 (3.7%) | 7 (7%) | 8 (2.6%) | 0.091 |
| Main symptom of illness onset |  |  |  | 0.122 |
| Fever | 319 (79.2%) | 87 (87%) | 232 (76.6%) |  |
| Cough | 41 (10.2%) | 4 (4%) | 37 (12.2%) |  |
| Fatigue | 14 (3.5%) | 2 (2%) | 12 (4.0%) |  |
| Sore throat | 8 (2.0%) | 0 (0) | 8 (2.6%) |  |
| Shortness of breath | 8 (2.0%) | 2 (2%) | 6 (2.0%) |  |
| Dyspnea | 5 (1.2%) | 4 (4%) | 1 (0.3%) |  |
| Diarrhea | 4 (1.0%) | 1 (1%) | 3 (1.0%) |  |
| Vomit | 2 (0.5%) | 0 (0) | 2 (0.7%) |  |
| Myalgia | 2 (0.5%) | 0 (0) | 2 (0.7%) |  |
| Days from onset to admission | 9 (7-12) | 9 (7-13) | 9 (6-11) | 0.188 |
| Abnormal imaging |  |  |  | 0.433 |
| Bilateral lungs | 376 (93.3%) | 95 (95%) | 281 (92.7%) |  |
| Single lung | 27 (6.7%) | 5 (5%) | 22 (7.3%) |  |

Age and days from onset to admission were presented as median (interquartile range) and compared with Mann-Whitney test between patients who died or recovered. Chi-squared test was conducted to compare other categorical variables between two groups.

Laboratory findings of patients with COVID-19 were listed in table 2 and table 3. Results of complete blood cell count test showed anemia in 220 (54.6%) and lymphopenia in 212 (52.6%) cases.

**Table 2.**
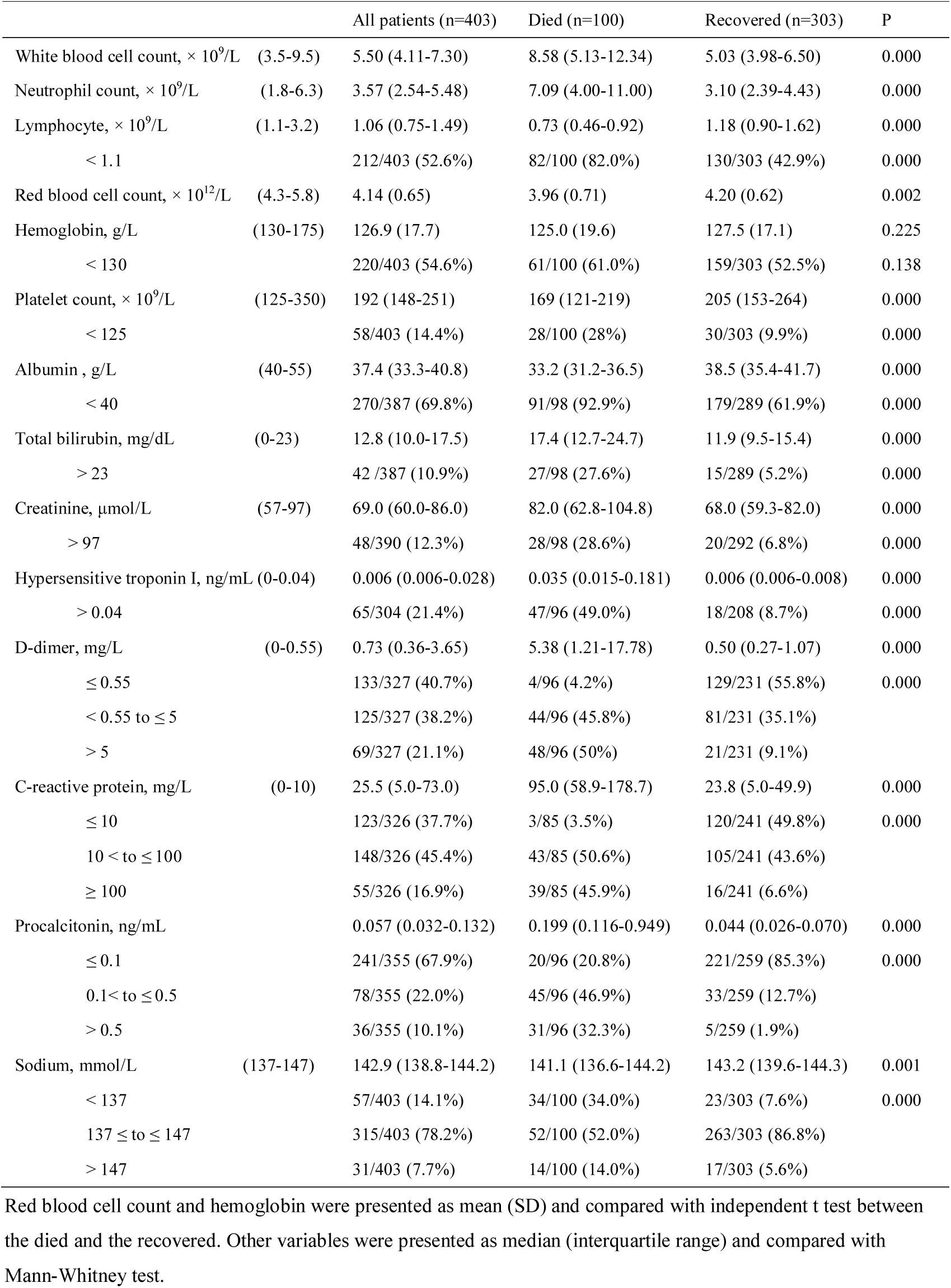
Laboratory findings of COVID-19 patients on admission according to clinical outcome.

**Table 3.** Clinical characteristics of COVID-19 patients according to disease severity on admission.

|  | Ordinary (n=198) | Severe or critical (n=205) | p |
| --- | --- | --- | --- |
| Age, years | 43 (33-56) | 66 (56-74) | 0.000 |
| Male gender | 77 (38.9%) | 116 (56.6%) | 0.000 |
| With chronic diseases | 56 (28.3%) | 119 (58.0%) | 0.000 |
| Days from onset to admission | 8 (5-11) | 10 (7-13) | 0.002 |
| White blood cell count, $\times 10^9/L$ | 4.91 (3.97-6.19) | 6.46 (4.47-8.93) | 0.000 |
| Neutrophil count, $\times 10^9/L$ | 2.94 (2.40-4.11) | 4.62 (2.97-7.97) | 0.000 |
| Lymphocyte count, $\times 10^9/L$ | 1.22 (0.94-1.70) | 0.90 (0.54-1.24) | 0.000 |
| < 1.1 | 74/198 (37.4%) | 138/205 (67.3%) | 0.000 |
| Red blood cell count, $\times 10^{12}/L$ | 4.25 (0.59) | 4.03 (0.68) | 0.000 |
| Hemoglobin, g/L | 128.8 (16.8) | 125.0 (18.4) | 0.031 |
| < 130 | 98/198 (49.5%) | 122/205 (59.5%) | 0.043 |
| Platelet count, $\times 10^9/L$ | 205 (154-261) | 179 (136-245) | 0.000 |
| < 125 | 16/198 (8.1%) | 42/205 (20.5%) | 0.000 |
| Albumin, g/L | 39.9 (37.3-42.2) | 34.7 (32.3-37.9) | 0.000 |
| < 40 | 94/184 (51.1%) | 176/203 (86.7%) | 0.000 |
| Total bilirubin, mg/dL | 11.6 (9.2-14.2) | 14.8 (10.9-20.8) | 0.000 |
| > 23 | 8/184 (4.3%) | 34/203 (16.7%) | 0.000 |
| Creatinine, $\mu\text{mol/L}$ | 68.0 (59.0-82.0) | 71.0 (61.0-88.0) | 0.028 |
| > 97 | 13/187 (7.0%) | 35/203 (17.2%) | 0.002 |
| Hypersensitive troponin I, ng/mL | 0.006 (0.006-0.006) | 0.012 (0.006-0.062) | 0.000 |
| > 0.04 | 13/127 (10.2%) | 52/177 (29.4%) | 0.000 |
| D-dimer, mg/L | 0.39 (0.23-0.83) | 1.22 (0.52-7.46) |  |
| $\leq 0.55$ | 80/135 (59.3%) | 53/192 (27.6%) | 0.000 |
| $0.55 < \text{to} \leq 5$ | 42/135 (31.1%) | 83/192 (43.2%) | 0.000 |
| > 5 | 13/135 (9.6%) | 56/192 (29.2%) |  |
| C-reactive protein, mg/L | 6.8 (5.0-25.7) | 59.7 (20.7-103.5) |  |
| $\leq 10$ | 88 (44.4%) | 33 (16.1%) | 0.000 |
| $10 < \text{to} \leq 100$ | 103 (52.0%) | 125 (61.0%) | 0.000 |
| $\geq 100$ | 7 (3.5%) | 47 (22.9%) | |
| Procalcitonin, ng/mL | 0.04 (0.02-0.06) | 0.10 (0.05-0.26) |  |
| $\leq 0.1$ | 142/161 (88.2%) | 99/194 (51.0%) | 0.000 |
| $0.1 < \text{to} \leq 0.5$ | 18/161 (11.2%) | 60/194 (30.9%) | 0.000 |
| > 0.5 | 1/161 (0.6%) | 35/194 (18.0%) |  |
| Sodium, mmol/L | 143.5 (142.2-145.0) | 141.2 (137.9-143.9) |  |
| < 137 | 10/198 (5.1%) | 47/205 (22.9%) | 0.000 |
| $\leq 137 \text{ to} \leq 147$ | 176/198 (88.9%) | 139/205 (67.8%) | 0.000 |
| > 147 | 12/198 (6.1%) | 19/205 (9.3%) |  |

Red blood cell count and hemoglobin were presented as mean (SD) and compared with independent t test. Other continuous variables were presented as median (interquartile range) and compared with Mann-Whitney test. Chi-squared test was conducted to compare categorical variables.

**Table 4.** Complications and treatments of COVID-19 patients.

|  | All patients (n=403) | Died (n=100) | Recovered (n=303) | P |
| --- | --- | --- | --- | --- |
| Complications |  |  |  |  |
| Acute respiratory distress syndrome | 143 (35.5%) | 90 (90.0%) | 53 (17.5%) | 0.000 |
| Acute kidney injury | 57 (14.1%) | 43 (43.0%) | 14 (4.6%) | 0.000 |
| Acute cardiac injury | 83 (20.6%) | 65 (65.0%) | 18 (5.9%) | 0.000 |
| Acute liver injury | 87 (21.6%) | 71 (71.0%) | 16 (5.3%) | 0.000 |
| Secondary infection | 77 (19.2%) | 58 (58.0%) | 19 (6.3%) | 0.000 |
| Treatment measures |  |  |  |  |
| High-flow nasal cannula oxygen therapy | 106 (26.3%) | 74 (74.0%) | 32 (10.6%) | 0.000 |
| Non-invasive mechanical ventilation | 56 (13.9%) | 48 (48.0%) | 8 (2.6%) | 0.000 |
| Invasive mechanical ventilation | 23 (5.7%) | 16 (16.0%) | 7 (2.3%) | 0.000 |
| Blood purification | 39 (9.7%) | 16 (16.0%) | 23 (7.6%) | 0.014 |
| Antiviral agents | 394 (97.8) | 96 (96.0%) | 298 (98.3%) | 0.323 |
| Abidol | 324 (80.4%) | 66 (66.0%) | 258 (85.1%) | 0.000 |
| Oseltamivir | 123 (30.5%) | 32 (21.0%) | 91 (30.0%) | 0.711 |
| Ribavirin | 81 (20.1%) | 26 (26.0%) | 55 (18.2%) | 0.089 |
| Ganciclovir | 53 (13.2%) | 17 (17.0%) | 36 (11.9%) | 0.189 |
| Recombinant interferon | 72 (17.9%) | 8 (8.0%) | 64 (21.1%) | 0.003 |
| Antibiotics | 349 (86.6%) | 99 (99.0%) | 250 (82.9%) | 0.000 |
| Anti-fungal agents | 14 (3.5%) | 11 (11.0%) | 3 (1.0%) | 0.000 |
| Glucocorticoids | 166 (41.2%) | 74 (74.0%) | 92 (30.4%) | 0.000 |
| Immunoglobulin | 198 (49.1%) | 66 (66.0%) | 132 (43.6%) | 0.000 |

Chi-squared test was conducted to compare categorical variables presented as number (%).

Non-survivors exhibited significantly higher white blood cell and neutrophil count, but lower red blood cell, lymphocyte and platelet count on admission than survivors. Two hundred and seventy out of 387 patients (69.8%) had serum albumin level below 40 g/L on admission. Increased serum total bilirubin and creatinine were observed in 10.9% (42/387) and 12.3% (48/390) patients, respectively. Non-survivors had much larger proportion of elevated total bilirubin (27.6% vs. 5.2%, p<0.001) and creatinine (28.6% vs. 6.8%, p<0.001) than survivors. Raised serum levels of hypersensitive troponin I (median 0.035 (0.015–0.181) ng/mL vs. 0.006 (0.006-0.008) ng/mL, p<0.001), D-dimer (median 5.38 (1.21–17.78) mg/L vs. 0.50 (0.27–1.07) mg/L, p<0.001), C-reactive protein (median 95.0 (58.9–178.7) mg/L vs. 23.8 (5.0–49.9) mg/L, p<0.001), and procalcitonin (median 0.199 (0.116–0.949) ng/mL vs. 0.044 (0.026–0.070) ng/mL, p<0.001) were observed in non-survivors compared to survivors. Abnormal serum sodium was observed in 21.8% (88) cases. Non-survivors showed lower serum sodium (median 141.1 (136.6–144.2) mmol/L vs. 143.2 (139.6–144.3) mmol/L, p=0.001) and higher incidence of abnormal serum sodium (48% vs. 13.2%, p<0.001) compared to survivors on admission. Two hundred and five (50.9%) cases were diagnosed as severe or critical illness, and 198 were ordinary illness on admission, according to the criteria issued by NHC^9^. Patients with severe or critical illness presented older age (median 66 (56–74) years vs. 43 (33–56) years, p <0.001), higher proportions of male gender (56.6% vs. 38.9%, p <0.001) or chronic diseases (58.0% vs. 28.3%, p <0.001), and more delayed hospitalization (median 10 (7–13) days vs. 8 (5–11) days, p <0.001) than those with ordinary illness. Decreased lymphocyte (median 0.90 (0.54–1.24) × 10^9^ /L vs. 1.22 (0.94–1.70) × 10^9^ /L, p <0.001) and platelet count (median 179 (136–245) × 10^9^ /L vs. 205 (154–261) × 10^9^ /L, p <0.001), and serum albumin level (median 34.7 (32.3–37.9) g/L vs. 39.9 (37.3–42.2) g/L, p <0.001) were observed in patients with severe or critical illness compared to those with ordinary disease. The incidence of increased serum total bilirubin (16.6% vs. 4.0%, p <0.001) and creatinine (16.7% vs. 4.3%, p =0.002) in patients with severe or critical disease were obviously higher than patients with ordinary disease. Patients with severe or critical illness showed markedly elevated serum level of hypersensitive troponin I (median 0.012 (0.006–0.062) ng/mL vs. 0.006 (0.006–0.006) ng/mL, p <0.001), D-dimer (median 1.22 (0.52–7.46) mg/L vs. 0.39 (0.23–0.83) mg/L, p <0.001), C-reactive protein (median 59.7 (20.7–103.5) mg/L vs. 6.8 (5.0–25.7) mg/L, p <0.001), and procalcitonin (median 0.10 (0.05–0.26) ng/mL vs. 0.04 (0.02–0.06) ng/mL, p <0.001) compared to those with ordinary illness. Decreased serum sodium appeared in patients with severe or critical illness compared to those with ordinary illness (median 141.2 (137.9–143.9) mmol/L vs. 143.5 (142.2–145.0) mmol/L, p<0.001). The proportion of abnormal serum sodium was significantly lower in patients with ordinary illness compared to those with severe or critical illness (12.2% vs. 32.2%, p<0.001).

Complications of acute organ injury and secondary infection were common in patients with COVID-19, especially in non-survivors. The incidence of acute respiratory distress syndrome (90% vs. 17.5%), acute liver injury (71% vs. 5.3%), cardiac injury (65% vs. 5.9%), kidney injury (43% vs. 4.6%), and secondary infection (58% vs. 6.3%) in patients who died was significantly higher than those who recovered (all p<0.001).

HFNC was used in 106 (26.3%) cases. NIMV and IMV were provided in 56 (13.9%) and 23 (5.7%) cases, respectively. Thirty nine (9.7%) patients received blood purification treatment. Antibiotics and anti-fungal agents were given to 349 (86.6%) and 14 (3.5%) cases respectively. Antiviral agents were given to 394 (97.8%) cases. Specifically, abidol was used in 324 (80.4%), oseltamivir in 123 (30.5%), ribavirin in 81 (20.1%), ganciclovir in 53 (13.2%), and recombinant interferon in 72 (17.9%). Almost half (49.1%) patients received immunoglobulin injection. One hundred and sixty-six (41.2%) patients received systemic application of glucocorticoids.

## Discussion

To our knowledge, this report presented the largest sample of COVID-19 patients with clinical outcome. Our results confirmed the findings by Chen that male gender, older age, and chronic disease contributed to death caused by SARS-CoV-2 infection.^7^ Patients with severe or critical illness on admission presented a more delay to admission, implying the effect of delayed admission on disease deterioration. The ongoing outbreak of this epidemic incurred a sharply increased patients in Wuhan, China, and most of them couldn’t be admitted to hospitals in time. Frequent visits to fever clinics, inappropriate home quarantine, and delayed hospitalization further aggravated the disease progression and transmission of the virus. Multiple homeostasis disturbances, including lymphopenia, anemia, hypoproteinemia and abnormal serum sodium were observed on admission, especially in patients with severe or critical illness. Many factors might contribute to these disturbances, such as eating less due to poor appetite for food, augment of organic consumption, and simultaneous gastrointestinal problems. Insufficient nutrient intake and augmented consumption might deteriorate tissue metabolism and damage one’s immunity to virus.

Our study showed that multiple organ damage, such as ARDS, acute kidney injury, acute cardiac injury, and acute liver injury, were common complications in patients with COVID-19. Previous studies have showed that angiotensin-convertion enzyme 2 (ACE2), the receptor for SARS-CoV-2, is expressed in organs including lung, heart, liver, and kidney. By binding to ACE2, SARS-CoV-2 can provide direct attacks to these organs.^10-12^ Serum CRP level on admission was evidently elevated in patients with COVID-19, especially in those with severe or critical illness, indicating excessive inflammatory stress, which was consistent with the elevated serum pro-inflammatory cytokines observed in COVID-19 patients.^13,14^ Meanwhile, serum levels of some laboratory indexes such as total bilirubin, creatinine and hypersensitive troponin I were increased in patients on admission. This trend was more obvious in patients with severe or critical illness, indicating signs of organ damage. Though the pathological mechanism of COVID-19 is yet to be defined, direct attacks from SARS-CoV-2 and organ damage caused by excessive inflammatory response (cytokines storm) might be responsible for the pathogenesis of disease progression.^14^ Secondary infection, another main complication in this study, appeared mostly in non-survivors, indicating an important booster of disease progression.

Comprehensive treatments were provided for patients in this study. No antiviral treatment for coronavirus infection has been proven to be effective, however, antiviral agents were applied in almost (97.8%) patients with COVID-19. The main agents used included abidol, oseltamivir, ribavirin, ganciclovir, and recombinant interferon. Remdesivir and chloroquine are considered two promising agents for SARS-CoV-2 infection,^15^ which were given to some patients in randomized, double-blind, placebo-controlled clinical trials. However, it’s hardly to assess the efficacy and safety of remdesivir and chloroquine until results of the trials are released. Short-term (3 to 5 days) application of glucocorticoids is suggested for severe patients with COVID-19 in the protocol issued by NHC.^9^ In this study, glucocorticoids were applied in 166 (41.2%) cases, most of whom were patients with severe or critical illness. However, glucocorticoids use was found to be correlated with increased mortality in SARS patients and delayed virus clearance.^16,17^ In this context, glucocorticoids are not routinely recommended in treatment of SARS-CoV-2 pneumonia.^18^ In this study, secondary infection appeared mostly in patients with glucocorticoids use and might contribute to death, which was indicated by its higher incidence in patients who died (58.0% vs. 6.3%, p<0.001). And so, glucocorticoids should be used more cautiously in patients with COVID-19.

High-flow nasal cannula oxygen therapy (HFNC) and mechanical ventilation are recommended for patients with ARDS.^19^ Non-invasive positive pressure ventilation (NIPPV) has advantage in improving oxygenation of patients with moderately or severely hypoxic respiratory failure (with oxygenation index PaO2/FiO2 100 to 150), however, it does not reduce rate of endotracheal intubation and ICU mortality.^20^ For patients with ARDS, invasive ventilation with strategy of pulmonary protection is strongly recommended.^21^ In this study, many patients with severe ARDS were not provided IMV treatment. Inadequate application of invasive ventilation in this study was mainly due to the shortage of intensive care unit (ICU) resource for so many critically ill patients accumulated during epidemic ongoing outbreak. The worry about aerosol transmission of virus during endotracheal intubation was possibly another reason for some medical staffs.^22^

There are some limitations in our study. First, current criteria for recovery are debatable since positive result of SARS-CoV-2 RNA detection appeared in some discharged patients after one to two weeks of continuous quarantine.^23^ Hence, some discharged patients may not truly recover. Second, the agents given to patients such as antibiotics, antivirus agents and anti-fungals, might exert toxicity to organs, which confounded judgment of organ damage by virus or excessive inflammatory stress. Besides, chronic diseases might also have effect on laboratory results on patients’ admission, especially in those with severe or critical illness. Third, inflammatory mediators, such as IL-1, IL-6, TNF-α, commonly being used to predict cytokines storm,^14^ were not included due to detection in just a small fraction of patients in this study. However, serum CRP level was detected for most patients in this study, which can also reflect inflammatory stress.

Since Dec 2019, SARS-CoV-2 has infected about fifty thousand patients and caused over two thousand deaths in Wuhan, China. With the strengthened support from central and local governments and national medical staffs, SARS-CoV-2 epidemic has been preliminary controlled in Wuhan, China at current. Many other countries such as Italy, Iran, and Korea, however, are showing an ongoing outbreak,^4^ which will inevitably bring about shortage of medical resources. To learn characteristics of patients with COVID-19 in Wuhan, China, during ongoing outbreak of SARS-CoV-2 epidemic, is helpful for others to make appropriate strategies and thus to reduce mortality of COVID-19 globally. In conclusion, findings of this study suggested multiple homeostasis disturbances in patients with severe or critical illness on admission, including lymphopenia, anemia, hypoproteinemia, abnormal serum sodium, and elevated inflammatory stress. Acute organ injuries were common in non-survivors, which might cause by SARS-CoV-2 attacks and excessive inflammatory response. Early supportive therapy should been provided to control disease progression and reduce the mortality of COVID-19.

## Data Availability

The data sets that support findings of the current study are available from the corresponding author on reasonable request.

## Acknowledgments

We thank all patients and their families involved in the study.

## Funding Sources

This study received no funding.

## Declaration of interests

All authors declare that they have no competing interests.

## Author Contributions

XL, LY and BZ had the idea for and designed the study and had full access to all data in the study and take responsibility for the integrity of the data and the accuracy of the data analysis. WZ, HX, BW and WY contributed to writing of the report. XL contributed to critical revision of the report. BW and TG contributed to the statistical analysis. All authors contributed to data acquisition, analysis, or interpretation, and reviewed and approved the final version.

## Notes

### Competing Interest Statement

The authors have declared no competing interest.

